# Multisite Real-World Validation of an Electronic Health Record-Integrated Generative Artificial Intelligence Tool for Venous Thromboembolism Risk Stratification

**DOI:** 10.64898/2026.06.17.26355819

**Authors:** Derek J. Baughman, Star Liu, Sangho Jee, Chase Young, Amy M. Knight, Andrew Davis, Srinivasan Yegnasubramanian, Allen Kachalia, Peter Najjar, James J. Whitbread, Luis Ahumada, Amy Chused, Elliott R. Haut, Brandyn D. Lau, Anirudh Sridharan, Michael Streiff, Khyzer B. Aziz

**Affiliations:** Biomedical Informatics and Data Science, Johns Hopkins School of Medicine, Baltimore, MD; Department of Medicine, Johns Hopkins School of Medicine, Baltimore, MD; Health IT, Johns Hopkins Medicine, Baltimore, MD; Department of Pulmonary and Critical Care Medicine, Johns Hopkins School of Medicine, Baltimore, MD; inHealth Precision Medicine, Johns Hopkins Medicine, Baltimore, MD; Sidney Kimmel Comprehensive Cancer Center, Johns Hopkins School of Medicine, Baltimore, MD; Armstrong Institute for Patient Safety and Quality, Johns Hopkins Medicine, Baltimore, MD; Department of Surgery, Johns Hopkins School of Medicine, Baltimore, MD; Johns Hopkins University School of Medicine, Baltimore, MD; Center for Pediatric Data Science and Analytic Methodology, Johns Hopkins All Children’s Hospital, St. Petersburg, FL; Department of Medicine, Johns Hopkins Sibley Memorial Hospital, Washington, DC; Department of Anesthesiology and Critical Care Medicine, Johns Hopkins School of Medicine, Baltimore, MD; Department of Emergency Medicine, Johns Hopkins School of Medicine, Baltimore, MD; Department of Health Policy and Management, Johns Hopkins Bloomberg School of Public Health, Baltimore, MD; Russell H. Morgan Department of Radiology and Radiological Science, Johns Hopkins School of Medicine, Baltimore, MD; Welch Center for Prevention, Epidemiology, and Clinical Research, Baltimore, MD; Department of Medicine, Johns Hopkins Howard County General Hospital, Columbia, MD; Department of Pediatrics, Johns Hopkins School of Medicine, Baltimore, MD

## Abstract

**Background:** Guiding risk-appropriate inpatient thromboprophylaxis requires venous thromboembolism (VTE) risk stratification; however, reliable risk determination remains inconsistent in routine care. Health systems increasingly pilot artificial intelligence (AI) tools, yet few studies demonstrate rigorous evaluation in the context of a learning health system (LHS). We evaluated the performance of a pilot electronic health record (EHR)-integrated generative AI (GenAI) system, inHealth General Reasoner (iHGR), for VTE risk stratification versus clinician order set classifications and physician-adjudicated chart review.

**Methods:** This multisite retrospective validation study included adult inpatient admissions at Johns Hopkins Medicine between June 21, 2025, and Dec 18, 2025 (checklist-based order set from June 21, 2025 – November 19, 2025, and clinician judgement-based order set from November 29 – December 18, 2025). From 758 eligible admissions, we randomly sampled 500 balanced by site and order set periods. iHGR and clinician-selected order set classifications were compared with the reference standard (RS). Primary outcomes were iHGR sensitivity and specificity. Secondary analyses compared the order sets with the same RS to evaluate workflow comparators and error patterns.

**Results:** iHGR achieved 81.8% sensitivity (95% CI 77.3–85.6) and 70.9% specificity (63.6–77.3). The checklist-based order set had 61.3% sensitivity (53.7–68.5) and 86.2% specificity (77.4–91.9). The clinician judgement-based order set had 78.1% sensitivity (71.3–83.7) and 65.4% specificity (54.3–75.0). False-negative iHGR classifications were associated with missed narrative risk factors.

**Conclusion:** iHGR showed higher sensitivity for VTE risk than checklist-based order sets and clinician judgement without introducing systematic bias. *In silico* evaluation of pilot AI systems within LHSs can identify clinically important performance trade-offs and implementation targets before operational scale-up. Narrative clinical data abstraction remained a key limitation, supporting the use of GenAI to support rather than supplant clinician judgement.

## INTRODUCTION

Venous thromboembolism (VTE) remains a leading cause of preventable morbidity and mortality among hospitalized patients and can be significantly mitigated by appropriate pharmacologic prophylaxis.^1^ Validated VTE risk assessment tools are widely recommended to guide prophylaxis decisions and have been used at the Johns Hopkins Hospital (JHH) for nearly two decades (see **Supplementary Appendix A**).^2^ For example, the internationally validated IMPROVE score^3^ incorporates clinical variables such as age, previous VTE, active cancer, thrombophilia, immobility, and intensive care unit admission for inpatient VTE risk assessment. Despite the availability of validated tools, implementation of VTE risk stratification remains inconsistent in routine care.^4^ In previous work at JHH, expert review agreed with admitting clinicians on overall VTE risk classification in only 65% of admissions, and appropriate prophylaxis was prescribed in 53% of cases.^5^ These findings highlight the difficulty of applying structured risk frameworks consistently at the point of care.^4^

### AI Opportunity & Literature Gap

Generative artificial intelligence (GenAI) and large language models (LLMs) have shown promise in extracting and synthesizing heterogeneous clinical information from electronic health records (EHRs). ^6^ To date, most clinical applications have focused on documentation, summarization, and information retrieval rather than support for guideline-based decision making.^7–9^ Simulaneously, health systems are increasingly piloting AI tools within routine workflows. This divergence between operational uptake and published evidence base is important: much of the literature remains centered on benchmarks, vignette, and experimental testing, with comparatively little evidence generated through methodologically rigorous study designs on how pilot systems perform in real care environments.

The clinical value of GenAI depends not only on the technical capability, but also on reliability, safety, and local performance under routine conditions. GenAI systems raise well-recognized concerns about hallucination, bias, inconsistent reasoning, and limited generalizability across settings.^10–13^ Yet, relatively few published studies describe rigorous *in silico* or silent evaluation of these systems within a learning-health-system framework for local validation, monitoring, and iterative refinement.^9^ Evidence is especially limited for EHR-integrated GenAI systems assessed on clinically meaningful decision tasks and compared directly with physician-adjudicated chart review.

### Study Rationale & Objectives

We evaluated a pilot EHR-integrated GenAI system, inHealth General Reasoner (iHGR), for inpatient VTE risk stratification using routine, real-world data from two hospitals. The primary objective was to assess iHGR performance against physician-adjudicated chart review as the reference standard. Secondary objectives were to compare iHGR with clinician-selected order set classifications across two workflow periods and to characterize error patterns in AI-assisted risk classification. As a learning health system evaluation, this study paired pilot deployment with retrospective *in silico* assessment to examine whether the system could classify risk and how such evaluation can inform local validation and iterative refinement before broader scale-up.

## METHODS

### Study Design

This multi-site retrospective validation study evaluated EHR data from 2 out of the 6 hospitals in the Johns Hopkins Health System (JHHS) - the Johns Hopkins Hospital, [JHH]) and Bayview Medical Center [BMC]) from June 21, 2025, to December 18, 2025. During this period, two order set workflows were used to classify VTE risk (**Supplementary Appendix B**). The checklist-based order set period, based on the Padua score,^14^ was from June 21, 2025, to November 19, 2025; November 20 - 28, 2025, was treated as a washout period; and the clinician judgement-based order set period, based on the IMPROVE score, was November 29, 2025, to December 18, 2025 (**Supplementary Appendix B Figure S1**). iHGR was applied across eligible encounters from both workflow periods. Patient-level classifications from the checklist-based order set, clinician judgement-based order set, and iHGR were compared with a physician-adjudicated chart review reference standard (**Figure 1**). The study was approved by the Johns Hopkins Medicine Institutional Review Board (IRB00531873) with consent waived for retrospective EHR review. We report this study according to the TRIPOD+AI guideline.^15^

**Figure 1:**
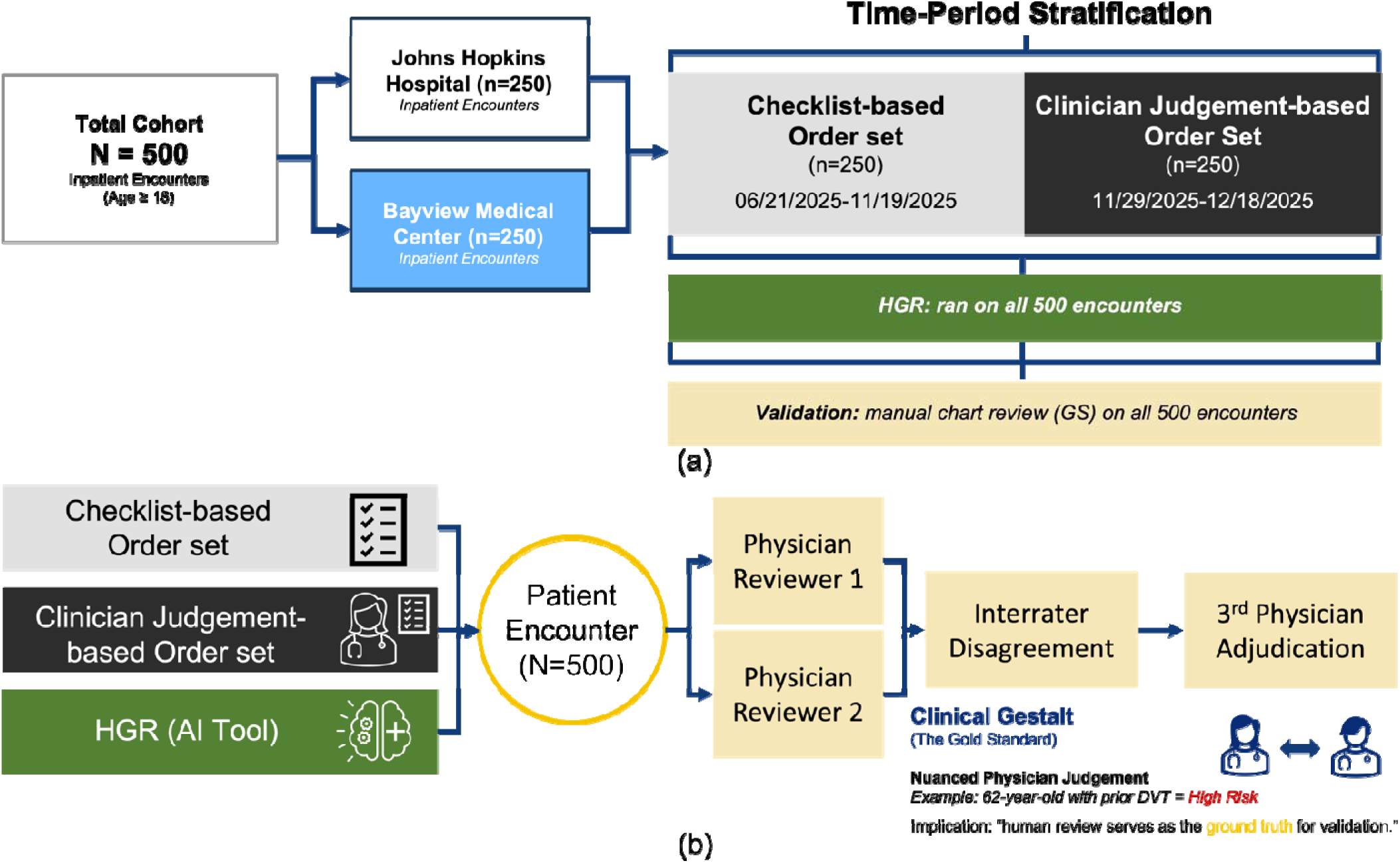
Study design and adjudication workflow. **(a)** Time-based stratification of the 500 adult inpatient encounters sampled from Johns Hopkins Hospital and Bayview Medical Center, with 250 encounters in the checklist-based order set period and 250 in the clinician judgement-based order set period. iHGR was applied across all encounters. **(b)** Comparison of VTE risk classification from the checklist-based order set, clin cian judgement-based order set, and iHGR against physician-adjudicated chart review. Two physicians independently reviewed each chart, with disagreements resolved by a third reviewer.

### Participants and Sampling

The study population included all adult inpatient admissions at JHH and BMC during the study period. Admissions without documented VTE order set were excluded. Each patient contributed one admission only, prioritizing the first admission in the clinician-judgement based order set period to account for small sample size. A stratified random sample of 500 patients was drawn from 75 unique clinicians across the two sites. To reduce clinician imbalance across workflow periods, clinicians with patients in both order set periods were prioritized. A maximum of eight patients per clinician per period was enforced to maintain balanced representation. After applying the inclusion and exclusion criteria, 758 unique hospital admissions were included as the denominator source from which the final analytic set (N=500) was sampled (**Figure 2**).

**Figure 2.**
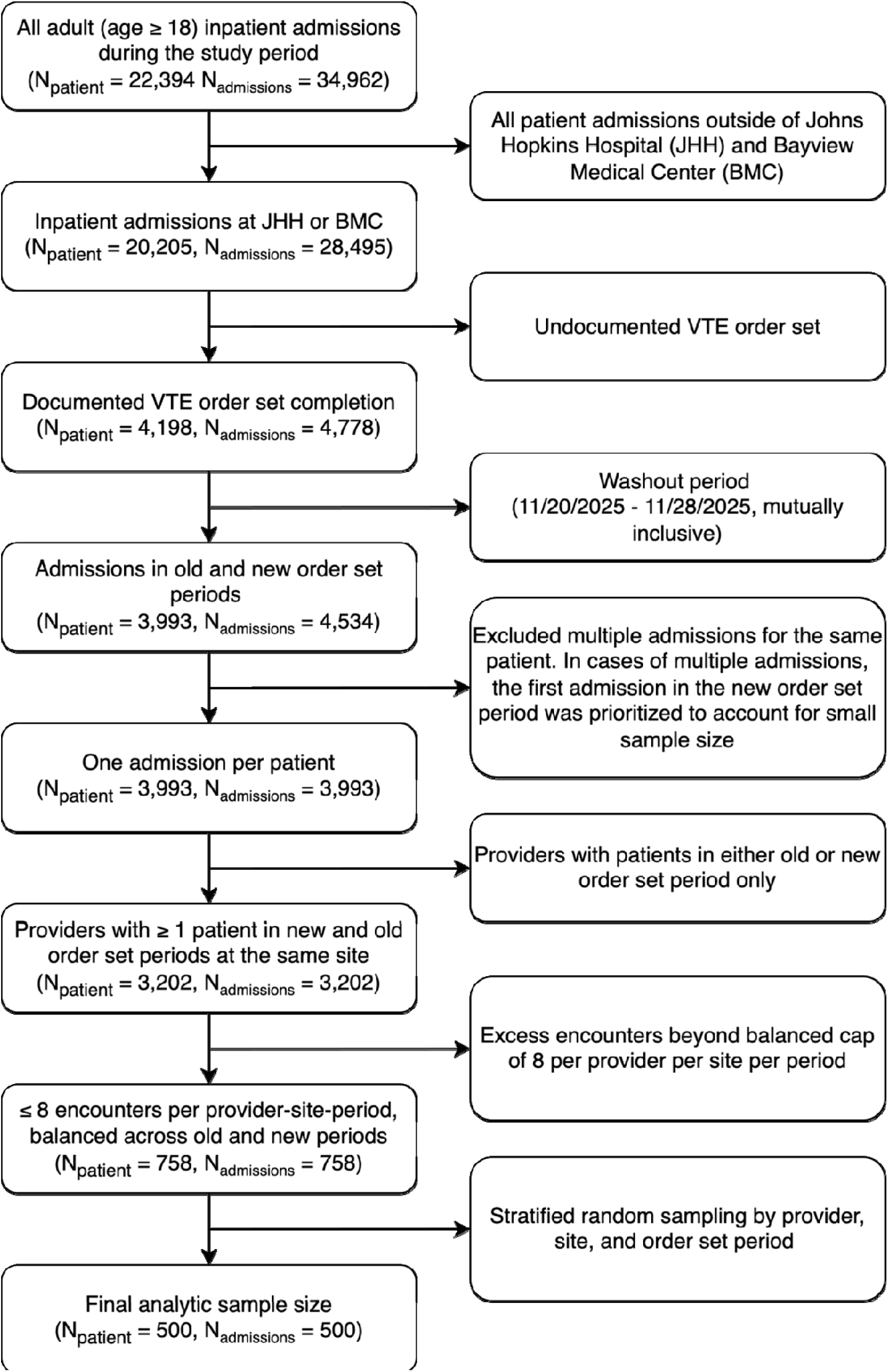
Consolidated Standards of Reporting Trials (CONSORT) diagram.

### Pilot AI System and Analytical Framework

iHGR was a pilot EHR-integrated GenAI system evaluated within routine care infrastructure during both order set periods. iHGR output was assessed retrospectively *in silico* rather than used as the determinant of clinical management. iHGR generated a binary VTE risk classification (standard vs. high-risk) from available structured EHR data and narrative clinical documentation (**Supplementary Appendix B**). iHGR did not explicitly calculate the IMPROVE score during inference. Instead, retrospective chart review and variable-level mapping to the IMPROVE framework were used as an analytic comparison process to standardize comparisons between iHGR outputs, clinician workflow decisions, and guideline-based risk variables. This distinction was important to the learning-health-system design: iHGR functioned as an independent reasoning system, whereas IMPROVE-based mapping was used *post hoc* to evaluate performance and identify targets for iterative refinement.

### Reference Standard

Two physicians independently reviewed all 500 charts in the EHR. Reviewers assessed IMPROVE-mapped risk factors and contraindications and then separately assigned an overall VTE risk classification of standard or high-risk based on full chart review and clinical judgement. Variable-level review was therefore distinguished from the reviewers’ final overall classification, allowing the study to compare structured guideline-based factors with broader clinical interpretation. Consistent with validated methods,^16^ discordances between the two reviewers were adjudicated by a third physician who was blinded to the initial chart-review results. The final adjudicated VTE risk classification served as the reference standard against which iHGR, the checklist-based order set, and the clinician judgement-based order set were compared. This design enabled comparison across three decision processes: structured questionnaire-based risk assessment (checklist-based order set), simplified clinician-entered workflow classification (clinician judgement-based order set), and AI-assisted reasoning from EHR data (iHGR).

### Outcomes

The primary outcome was iHGR performance in VTE risk classification compared with the reference standard. Secondary outcomes were the performance of the checklist-based and clinician judgement-based order sets against the same reference standard, and the characterization of misclassification patterns relevant to local learning and system refinement.

### Statistical Analysis

The analytic sample size was determined by balancing the detection of minimum 8-10% improvement in sensitivity (80% power and α=0.05) against the practical constraints of performing physician chart reviews. The final sample size was 500 patients. Physician interrater reliability was measured using percent agreement, positive and negative agreement, McNemar’s test, Cohen’s kappa (κ); Prevalence Adjusted Bias Adjusted Kappa (PABAK), and Gwet’s AC_1_.^17–19^

For descriptive statistics, we compared age, sex, race, ethnicity, marital status, employment, language, Charlson Comorbidity Index score, length of stay (LOS), and mortality distributions. The Kruskal-Wallis test was performed for differences in nonparametric continuous variables, and the chi-square test was performed for difference in proportions.

Using the physician-adjudicated review as the reference standard, performance measures included sensitivity, specificity, accuracy, and Cohen’s κ. 95% confidence intervals (CI) were calculated using the Wilson score method. Classifications were categorized as true positives (TP), false positives (FP), false negatives (FN), and true negatives (TN). TPs were truly high-risk patients. FPs were standard risk patients incorrectly flagged as high-risk. FNs were high-risk patients incorrectly flagged as standard risk. TNs were truly standard risk patients. All analyses were conducted in Python 3.12.3. Because the study was designed as a learning-health-system evaluation of a pilot deployment, we analyzed error patterns to inform local workflow and future system optimization.

## RESULTS

### Cohort and Sampling Balance

A total of 75 clinicians contributed to the analytic sample (44 at BMC and 31 at JHH). Each matched clinician contributed a mean of 3.5 patients in the checklist-based order set period and 3.6 patients in the clinician judgement-based order set period (median of 3 in both periods). Of these, 67 clinicians (89%) had patients in both workflow periods, permitting within-clinician comparison across the workflow change (See **Supplementary Appendix C Table S1 and Figure S1).**

Compared with the 3,202 admissions remaining after clinician matching, the analytic sample did not differ significantly in age, sex, race, ethnicity, marital status, employment status, language, LOS, and mortality (**Table 1**). The median age was 63.0 years; 55% were female; and the median LOS was 4 days. Charlson Comorbidity Index distributions differed, although the interquartile ranges overlapped. Compared with BMC, the JHH subgroup differed in race and marital status distributions, but was otherwise similar in Charlson Comorbidity Index score, LOS, and mortality. (**Supplementary Appendix C Table S2**).

**Table 1.** Patient demographics among all adult inpatient admissions, post clinician matching and balancing, and the final stratified randomly sampled 500 patients.

| **Patients (N)** | **Post Clinician Matching**<br>**3202** | **Analytic Sample**<br>**500** | **P-Value** |
| --- | --- | --- | --- |
| **Age, median [Q1,Q3]** | 63.5 [48.2,75.1] | 63.0 [46.8,75.6] | 0.523 |
| **Sex, n (%)** |  |  |  |
| Female | 1676 (52.3) | 275 (55.0) | 0.505 |
| Male | 1525 (47.6) | 225 (45.0) |  |
| Other/Unknown | 1 (0.0) |  |  |
| **Race, n (%)** |  |  |  |
| White | 1738 (54.3) | 257 (51.4) | 0.768 |
| Black | 1211 (37.8) | 201 (40.2) |  |
| Other/Multiple | 185 (5.8) | 29 (5.8) |  |
| Asian | 64 (2.0) | 12 (2.4) |  |
| Unknown/Not Disclosed | 4 (0.1) | 1 (0.2) |  |
| **Ethnicity, n (%)** |  |  |  |
| Not Hispanic/Latino | 2413 (75.4) | 369 (73.8) | 0.683 |
| Other/Unknown | 669 (20.9) | 113 (22.6) |  |
| Hispanic/Latino | 120 (3.7) | 18 (3.6) |  |
| **Marital Status, n (%)** |  |  |  |
| Married | 899 (28.1) | 149 (29.8) | 0.864 |
| Single | 1330 (41.5) | 208 (41.6) |  |
| Widowed | 457 (14.3) | 69 (13.8) |  |
| Divorced | 353 (11.0) | 53 (10.6) |  |
| Other/Unknown | 163 (5.1) | 21 (4.2) |  |
| **Employment Status, n (%)** |  |  |  |
| Retired | 1216 (38.0) | 195 (39.0) | 0.365 |
| Employed | 606 (18.9) | 103 (20.6) |  |
| Not Employed | 909 (28.4) | 131 (26.2) |  |
| Disabled | 423 (13.2) | 59 (11.8) |  |
| Other/Unknown | 48 (1.5) | 12 (2.4) |  |
| **Language, n (%)** |  |  |  |
| English | 3062 (95.6) | 474 (94.8) | 0.701 |
| Spanish | 106 (3.3) | 20 (4.0) |  |
| Other | 34 (1.1) | 6 (1.2) |  |
| **Charlson Comorbidity Score, median [Q1,Q3]** | 1.0 [0.0,3.0] | 2.0 [0.0,3.0] | 0.012 |
| **Length of Stay (days), median [Q1,Q3]** | 4.0 [2.0,8.0] | 4.0 [2.0,8.0] | 0.375 |
| **Mortality, n (%)** | 211 (6.6) | 30 (6.0) | 0.689 |

### Reference Standard Reliability

Among the 500 cases, the two primary reviewers agreed in 378 (75.6%) cases (**Supplementary Appendix C Table S3**). Cohen’s κ was 0.488, PABAK was 0.512, Gwet’s AC1 was 0.534, and the Krippendorff’s Alpha was 0.489. Among the 122 discordant cases, adjudication produced 74 high-risk and 48 standard-risk classifications. Inter-rater agreement on high-risk was 0.799 and standard risk was 0.689. Findings indicate expected variability in clinical review and support the adjudicated chart review as a reference standard for learning-health-system evaluation of the pilot AI system.

### Primary iHGR Performance

The adjudicated chart review reference standard identified 335 (67%) high-risk and 165 (33%) standard risk cases (**Table 2**). Relative to this reference standard, iHGR yielded 274 TPs, 48 FPs, 61 FNs, and 117 TNs. Overall, iHGR achieved 81.8% (77.3-85.6) sensitivity, 70.9% (63.6-77.3) specificity, and 78.2% (74.4-81.6) accuracy, with moderate agreement with adjudicated review (κ=0.517, 95% CI 0.436–0.597). iHGR showed higher sensitivity at BMC and higher specificity at JHH, although site-specific CIs overlapped.

**Table 2.** iHGR primary outcome performance compared to the reference standard (RS) chart review.

| **Primary Outcomes** | **N** | **TP** | **FP** | **FN** | **TN** | **Sensitivity (95% CI)** | **Specificity (95% CI)** | **Accuracy (95% CI)** |
| --- | --- | --- | --- | --- | --- | --- | --- | --- |
| **iHGR vs RS** |  |  |  |  |  |  |  |  |
| Overall | 500 | 274 | 48 | 61 | 117 | **0.818 (0.773-0.856)** | **0.709 (0.636-0.773)** | **0.782 (0.744-0.816)** |
| JHH | 250 | 141 | 18 | 36 | 55 | 0.797 (0.731-0.849) | 0.753 (0.644-0.838) | 0.784 (0.729-0.831) |
| BMC | 250 | 133 | 30 | 25 | 62 | 0.842 (0.777-0.890) | 0.674 (0.573-0.761) | 0.780 (0.725-0.827) |
| **Secondary Outcomes** | **N** | **TP** | **FP** | **FN** | **TN** | **Sensitivity (95% CI)** | **Specificity (95% CI)** | **Accuracy (95% CI)** |
| **Checklist-based OS vs RS** |  |  |  |  |  |  |  |  |
| Overall | 250 | 100 | 12 | 63 | 75 | **0.613 (0.537-0.685)** | **0.862 (0.774-0.919)** | **0.700 (0.641-0.753)** |
| JHH | 125 | 54 | 5 | 29 | 37 | 0.651 (0.543-0.744) | 0.881 (0.750-0.948) | 0.728 (0.644-0.798) |
| BMC | 125 | 46 | 7 | 34 | 38 | 0.575 (0.466-0.677) | 0.844 (0.712-0.923) | 0.672 (0.586-0.748) |
| **Clinician judgement-based OS (Excluding anticoagulated) vs RS** |  |  |  |  |  |  |  |  |
| Overall | 197 | 83 | 26 | 37 | 51 | **0.692 (0.604-0.767)** | **0.662 (0.551-0.758)** | **0.680 (0.612-0.741)** |
| JHH | 104 | 53 | 10 | 20 | 21 | 0.726 (0.614-0.815) | 0.677 (0.501-0.814) | 0.712 (0.618-0.790) |
| BMC | 93 | 30 | 16 | 17 | 30 | 0.638 (0.495-0.760) | 0.652 (0.508-0.773) | 0.645 (0.544-0.735) |
| **Clinician judgement-based OS (Anticoagulated mapped to high-risk) vs RS** |  |  |  |  |  |  |  |  |
| Overall | 247 | 132 | 27 | 37 | 51 | **0.781 (0.713-0.837)** | **0.654 (0.543-0.750)** | **0.741 (0.683-0.792)** |
| JHH | 124 | 73 | 10 | 20 | 21 | 0.785 (0.691-0.856) | 0.677 (0.501-0.814) | 0.758 (0.676-0.825) |
| BMC | 123 | 59 | 17 | 17 | 30 | 0.776 (0.671-0.855) | 0.638 (0.495-0.760) | 0.724 (0.639-0.795) |

iHGR: inHealth General Reasoner. OS: Order set. RS: Reference standard chart review. TP: True positive. FP: False positive. FN: False negative. TN: True negative.

### Comparison with Clinician Workflows

Compared with clinician workflows, the checklist-based order set had lower sensitivity but higher specificity than iHGR, with sensitivity of 61.3% (95% CI 53.7–68.5) and specificity of 86.2% (77.4–91.9). The clinician judgement-based order set, with anticoagulated status mapped to high-risk, showed sensitivity of 78.1% (71.3–83.7) and specificity of 65.4% (54.3–75.0), moving closer to iHGR in sensitivity while remaining lower in agreement with the reference standard. Results suggest that workflow design materially influenced clinician-entered risk classification and provided an appropriate comparator context for local evaluation of the pilot AI system.

### Error Patterns

iHGR’s FN classifications were most often associated with missed narrative or context-dependent risk factors were, particularly immobility (22 [36.1%] of 61), previous DVT or PE (17 [27.9%]), and thrombophilia (15 [24.6%]), **Table 3**. Across the pilot deployment evaluation, these error patterns suggested that iHGR performed more reliably when key factors were readily available in structured EHR data and less reliably when classification depended on interpretation of free-text clinical documentation. This pattern was the most actionable output of the *in silico* evaluation for subsequent local refinement.

**Table 3.** Reference standard (RS) chart review variables stratified by iHGR classification groups.

| Variables | RS | iHGR Risk Classification |  |  |  |
| --- | --- | --- | --- | --- | --- |
|  | (N=500) | FN (N=61) | FP (N=48) | TN (N=117) | TP (N=274) |
| Age > 60, n (%) | 280 (56.0) | 5 (8.2) | 42 (87.5) | 3 (2.6) | 230 (83.9) |
| History of DVT or PE, n (%) | 148 (29.6) | 17 (27.9) | 2 (4.2) | 0 (0.0) | 129 (47.1) |
| ICU Care, n (%) | 36 (7.2) | 10 (16.4) | 0 (0.0) | 0 (0.0) | 26 (9.5) |
| Active Cancer, n (%) | 101 (20.2) | 10 (16.4) | 4 (8.3) | 1 (0.9) | 86 (31.4) |
| Thrombophilia, n (%) | 46 (9.2) | 15 (24.6) | 0 (0.0) | 2 (1.7) | 29 (10.6) |
| Acute Lower Limb Paralysis, n (%) | 28 (5.6) | 9 (14.8) | 0 (0.0) | 0 (0.0) | 19 (6.9) |
| Immobility/Bedrest, n (%) | 110 (22.0) | 22 (36.1) | 6 (12.5) | 3 (2.6) | 79 (28.8) |
| High Bleeding Risk, n (%) | 93 (18.6) | 14 (23.0) | 3 (6.2) | 14 (12.0) | 62 (22.6) |
| On Anticoagulants, n (%) | 136 (27.2) | 11 (18.0) | 1 (2.1) | 2 (1.7) | 122 (44.5) |
| Liver Disease, n (%) | 137 (27.4) | 21 (34.4) | 11 (22.9) | 32 (27.4) | 73 (26.6) |
| In Hospice Care, n (%) | 5 (1.0) | 0 (0.0) | 0 (0.0) | 0 (0.0) | 5 (1.8) |
| Thrombolytics in the last 24 hours, n (%) | 0 (0.0) | 0 (0.0) | 0 (0.0) | 0 (0.0) | 0 (0.0) |
| aPTT (seconds), mean (SD) | 32.3 (12.9) | 39.4 (22.5) | 26.5 (2.8) | 27.1 (3.2) | 33.0 (12.2) |
| INR, mean (SD) | 1.2 (0.4) | 1.3 (0.3) | 1.2 (0.4) | 1.1 (0.1) | 1.2 (0.5) |
| Platelet Count (K/mcL), mean (SD) | 238.7 (110.6) | 260.1 (144.0) | 262.3 (112.9) | 242.1 (86.1) | 228.3 (110.0) |

iHGR: inHealth General Reasoner. RS: Reference standard chart review. TP: True positive. FP: False positive. FN: False negative. TN: True negative. N: No, indicating the element was deemed false by the RS. Y: Yes, indicating the element was deemed true by the RS.

Further analysis uncovered the sources of risk factors at the time of admission that the iHGR used for risk classification (**Supplementary Appendix C Table S4**). Among the 48 FPs that should have been standard risk, the iHGR found age greater than 60 as the only risk factor in 38 (79.2%) cases. 4 FPs had at least one additional risk factor. 5 FPs were due to history of VTE.

## DISCUSSION

In this multisite, retrospective validation study, iHGR showed higher sensitivity than clinician order set workflow for identifying patients at high VTE risk, with moderate agreement relative to physician-adjudicated chart review. Framed as a learning-health-system evaluation, the study paired pilot deployment with retrospective *in silico* assessment to support local validation and iterative refinement before broader scale-up. The principal source of residual error was incomplete capture of narrative and context-dependent risk factors, suggesting that current GenAI systems can perform well when clinically relevant information is explicit and accessible, but remain limited when classification depends on inference from free-text documentation.

This study addressed a substantial evidence gap between technical AI development and local clinical evaluation under real EHR conditions.^9^ Although AI tools are increasingly piloted or operationalized in health systems, relatively few published studies describe rigorous *in silico* or silent evaluation as part of a learning-health-system approach to local validation, monitoring, and iterative refinement.^9,20,21^ In that context, the contribution of this study is not simply AI system workflow integration, but also an evaluation of pilot deployment against both clinically meaningful workflow comparators and a physician-adjudicated reference standard. Benchmark performance or simulated testing alone does not establish how a system behaves when applied to routine hospital data, local documentation practices, and real operational alternatives.^22^

Prior AI studies on VTE risk have largely focused on two specific tasks: prediction of future VTE events and identification of documented VTE events or related concepts within the EHR.^23,24^ By contrast, prophylaxis-relevant risk classification at admission - the task most directly linked to routine thromboprophylaxis decisions - remains less well studied, particularly for GenAI systems embedded within the EHR.^23–26^ Our study therefore extends the literature by evaluating an AI system designed to support a clinically actionable decision task, rather than risk prediction alone. It also differs from previous literature in that iHGR functioned as an independent reasoning system using structured and narrative data, whereas the IMPROVE framework served as an analytic comparison framework rather than the mechanism by which the AI generated its classification.

Direct comparison of AI-based VTE risk classification with physician-adjudicated chart review is largely absent from the literature.^24,25^ This matters because clinician disagreement in VTE assessment is common: at Johns Hopkins, concordance between frontline physicians and expert hematology review has previously been limited^5^ and inter-rater agreement for VTE risk assessment varies across settings.^27^ Physicians agree less with computer-calculated VTE risk scores, with reported discordance rates of ∼45%.^28^ Reported inter-rater agreement for clinician VTE risk assessment also varies substantially, ranging from κ≈0.1–0.5 depending on the patient population and risk definition.^5,27^ Against that background, the moderate reviewer agreement observed in our chart review was expected, and agreement between iHGR and the adjudicated reference standard should be interpreted within the known variability of clinical judgement.^5,28^ The use of adjudication was therefore a key feature of the study design, allowing the AI system and clinician workflows to be assessed against a clinically grounded reference process rather than an unreviewed proxy label.

The error patterns observed in this study informs clinical implementation. iHGR performed best when relevant risk factors were explicit and available in structured EHR data. Conversely, FN classifications were concentrated in factors such as immobility, previous DVT or PE, thrombophilia, and other concepts that often-required interpretation of free-text documentation or broader clinical context (**Supplementary Appendix B Figure S2**). These findings suggest that current GenAI systems can consolidate dispersed but explicit information effectively but remain less reliable when clinical reasoning depends on subtle narrative cues, incomplete documentation, or contextual inference. Therefore, error analysis may be more informative than headline performance metrics alone for judging implementation readiness.

The comparison with clinician workflows also has practical implications. First, AI can curate and streamline the review process by integrating relevant structured and narrative data, thereby reducing the need for repeated micro-decisions during routine risk assessment.^5,29^ Second, reducing micro-decision-making help preserve clinician judgement for cases in which risk classification depends on incomplete context, inference, or trade-offs that are not fully represented in structured fields. In this study, iHGR appeared most useful not as a replacement for clinician assessment, but as a system that could support more consistent identification of high-risk patients while leaving final interpretation in context-sensitive cases to clinicians.

Study findings should be interpreted considering the interaction between model performance and workflow design. iHGR achieved higher sensitivity than both clinician order set workflows, whereas the checklist-based order set retained higher specificity. The newer workflow moved closer to iHGR in sensitivity when anticoagulated status was mapped to high-risk, suggesting that observed differences in classification performance reflected not only the AI system itself, but also the design of the comparator workflows. This observation reinforces that evaluation of clinical AI should account for the surrounding decision architecture rather than treating model output as independent from the workflow in which it is used.^30^

This study has several strengths. First, we evaluated the system in a balanced sample drawn from multiple hospitals within the same health system, allowing assessment across heterogeneous care settings. Second, physician-adjudicated chart review provided a clinically grounded basis for comparison. Third, the study compared AI output with two clinician workflow configurations, enabling interpretation of AI performance relative to real operational alternatives rather than in isolation. Finally, detailed variable-level and error review identified discordance sources that inform future AI system and clinical workflow refinements.

### Limitations

This study should be interpreted in the context of its learning-health-system design for the evaluation of pilot AI deployments. The analysis was retrospective and *in silico*, designed to evaluate model performance under controlled conditions. The effects of live AI-assisted decision support on clinician behavior, prophylaxis prescribing, or patient outcomes were outside the scope of this study. Similarly, VTE risk was assessed at admission and therefore did not capture dynamic changes in risk during hospitalization. Because the consequences of FN and FP classifications are asymmetric, prospective implementation studies are needed to evaluate how model performance changes as clinical risk evolves. Future work should relate model errors to downstream patient-centered outcomes, evaluating whether classification errors translate into prophylaxis decisions, incident VTE, bleeding complications, and alert burden.

## CONCLUSION

Study findings suggest that a pilot EHR-integrated GenAI system can support inpatient VTE risk stratification with greater sensitivity than clinician order set workflows, although performance remains constrained by incomplete capture and interpretation of narrative clinical data. This work set the stage for further prospective evaluation of GenAI as an adjunct to clinician judgement and highlight retrospective *in silico* assessment as a valuable component of learning-health-system governance for early-stage AI deployment.

## AKNOWLEDGMENTS

### Funding

Authors D.B. and S.L. were supported by the National Library of Medicine T15 Training Grant (NLM 5T15LM013979) through the Johns Hopkins Biomedical Informatics and Data Science program. The funder of the study had no role in study design, data collection, data analysis, data interpretation, or writing of the manuscript. K.A., C.Y., and S.Y. were funded by Johns Hopkins inHealth Precision Medicine.

### Potential Conflict of Interests

No authors have C.O.I. to report.

### Author Contributions

Conceptualization & Methodology: K.A., D.B., S.J., S.L.

Formal Analysis, Investigation, Data Curation: K.A., D.B., S.J., S.L.

Writing—Original Draft: K.A., D.B., S.J., S.L.

Writing—Review & Editing: all authors contributed.

Supervision: K.A., D.B.

### Data Sharing Statement

De-identified participant data and data dictionary will be made available to researchers who provide a methodologically sound proposal, beginning 6 months after publication, for a period of 5 years. Proposals should be directed to. Data requestors will need to sign a data access agreement.

## Notes

### Competing Interest Statement

The authors have declared no competing interest.

### Author Declarations

The study was approved by the Johns Hopkins Medicine Institutional Review Board (IRB00531873) with consent waived for retrospective electronic health record review.

### Summary of Updates

One coauthor was accidently left out of the first submission.

## REFERENCES

1. Martin KA, Sparks AD, Wilkinson K, Packer R, Poston JN, Terrell DR, et al. Impact of hospital-acquired venous thromboembolism on surviving a medical admission: findings from the Medical Inpatient Thrombosis and Hemostasis Study. J Thromb Haemost JTH. 2026 Jan;24(1):227–33. doi:10.1016/j.jtha.2025.09.033 PubMed PMID: 41135659; PubMed Central PMCID: PMC12673607.

2. Henke PK, Kahn SR, Pannucci CJ, Secemksy EA, Evans NS, Khorana AA, et al. Call to Action to Prevent Venous Thromboembolism in Hospitalized Patients: A Policy Statement From the American Heart Association. Circulation. 2020 Jun 16;141(24):e914–31. doi:10.1161/CIR.0000000000000769

3. Rosenberg D, Eichorn A, Alarcon M, McCullagh L, McGinn T, Spyropoulos AC. External validation of the risk assessment model of the International Medical Prevention Registry on Venous Thromboembolism (IMPROVE) for medical patients in a tertiary health system. J Am Heart Assoc. 2014 Nov 17;3(6):e001152. doi:10.1161/JAHA.114.001152 PubMed PMID: 25404191; PubMed Central PMCID: PMC4338701.

4. Häfliger E, Kopp B, Darbellay Farhoumand P, Choffat D, Rossel JB, Reny JL, et al. Risk Assessment Models for Venous Thromboembolism in Medical Inpatients. JAMA Netw Open. 2024 May 10;7(5):e249980. doi:10.1001/jamanetworkopen.2024.9980

5. Lau BD, Bhave A, Yui JC, Naik R, Dane KE, Lindsley J, et al. Inaccuracies of venous thromboembolism risk assessment and prevention practices among medically ill patients. Blood Adv. 2025 Aug 12;9(15):3929–36. doi:10.1182/bloodadvances.2024015306 PubMed PMID: 40408310; PubMed Central PMCID: PMC12336698.

6. Shao C, Snyder D, Li C, Gu B, Ngan K, Yang CT, et al. Scalable medication extraction and discontinuation identification from electronic health records using large language models. J Clin Epidemiol. 2026 Jan;189:112049. doi:10.1016/j.jclinepi.2025.112049 PubMed PMID: 41232578; PubMed Central PMCID: PMC12714491.

7. Maddox TM, Embí P, Gerhart J, Goldsack J, Parikh RB, Sarich TC. Generative AI in Medicine — Evaluating Progress and Challenges. N Engl J Med. 2025 Jun 25;392(24):2479–83. doi:10.1056/NEJMsb2503956

8. Zhang Z, Momeni Nezhad MJ, Bagher Hosseini SM, Zolnour A, Zonour Z, Hosseini SM, et al. Advancing healthcare with large language models: A scoping review of applications and future directions. Int J Med Inf. 2026 Mar 15;208:106231. doi:10.1016/j.ijmedinf.2025.106231 PubMed PMID: 41443123.

9. Bedi S, Liu Y, Orr-Ewing L, Dash D, Koyejo S, Callahan A, et al. Testing and Evaluation of Health Care Applications of Large Language Models: A Systematic Review. JAMA. 2025 Jan 28;333(4):319–28. doi:10.1001/jama.2024.21700

10. Naliyatthaliyazchayil P, Muthyala R, Gichoya JW, Purkayastha S. Evaluating the Reasoning Capabilities of Large Language Models for Medical Coding and Hospital Readmission Risk Stratification: Zero-Shot Prompting Approach. J Med Internet Res. 2025 Jul 30;27:e74142. doi:10.2196/74142 PubMed PMID: 40737604; PubMed Central PMCID: PMC12310144.

11. Lee H, Xiong C, Baughman D, Dun C, Tong J, Martin B, et al. A multidimensional hierarchical framework for sources of bias in real-world healthcare evidence: a scoping review. J Biomed Inform. 2026 Feb;174:104989. doi:10.1016/j.jbi.2026.104989 PubMed PMID: 41571173.

12. Rouzrokh P, Khosravi B, Faghani S, Moassefi M, Shariatnia MM, Rouzrokh P, et al. A Current Review of Generative AI in Medicine: Core Concepts, Applications, and Current Limitations. Curr Rev Musculoskelet Med. 2025 Jul;18(7):246–66. doi:10.1007/s12178-025-09961-y PubMed PMID: 40304941; PubMed Central PMCID: PMC12185825.

13. Deng R, Martin G, Wang T, Zhang G, Liu Y, Weng C, et al. CPGPrompt: translating clinical guidelines into large language model-executable decision support. J Am Med Inform Assoc JAMIA. 2026 Feb 26;ocag026. doi:10.1093/jamia/ocag026 PubMed PMID: 41746783.

14. Barbar S, Noventa F, Rossetto V, Ferrari A, Brandolin B, Perlati M, et al. A risk assessment model for the identification of hospitalized medical patients at risk for venous thromboembolism: the Padua Prediction Score. J Thromb Haemost JTH. 2010 Nov;8(11):2450–7. doi:10.1111/j.1538-7836.2010.04044.x PubMed PMID: 20738765.

15. Collins GS, Moons KGM, Dhiman P, Riley RD, Beam AL, Calster BV, et al. TRIPOD+AI statement: updated guidance for reporting clinical prediction models that use regression or machine learning methods. BMJ. 2024 Apr 16;385:e078378. doi:10.1136/bmj-2023-078378 PubMed PMID: 38626948.

16. Kellerhuis BE, Jenniskens K, Kusters MPT, Schuit E, Hooft L, Moons KGM, et al. Expert panel as reference standard procedure in diagnostic accuracy studies: a systematic scoping review and methodological guidance. Diagn Progn Res. 2025 May 13;9:12. doi:10.1186/s41512-025-00195-7 PubMed PMID: 40355966; PubMed Central PMCID: PMC12070646.

17. Chen G, Faris P, Hemmelgarn B, Walker RL, Quan H. Measuring agreement of administrative data with chart data using prevalence unadjusted and adjusted kappa. BMC Med Res Methodol. 2009 Jan 21;9:5. doi:10.1186/1471-2288-9-5 PubMed PMID: 19159474; PubMed Central PMCID: PMC2636838.

18. Wongpakaran N, Wongpakaran T, Wedding D, Gwet KL. A comparison of Cohen’s Kappa and Gwet’s AC1 when calculating inter-rater reliability coefficients: a study conducted with personality disorder samples. BMC Med Res Methodol. 2013 Apr 29;13:61. doi:10.1186/1471-2288-13-61 PubMed PMID: 23627889; PubMed Central PMCID: PMC3643869.

19. Cicchetti DV, Feinstein AR. High agreement but low kappa: II. Resolving the paradoxes. J Clin Epidemiol. 1990;43(6):551–8. doi:10.1016/0895-4356(90)90159-m PubMed PMID: 2189948.

20. Sahni NR, Carrus B. Artificial Intelligence in U.S. Health Care Delivery. N Engl J Med. 2023 Jul 26;389(4):348–58. doi:10.1056/NEJMra2204673

21. Cabitza F, Campagner A, Balsano C. Bridging the “last mile” gap between AI implementation and operation: “data awareness” that matters. Ann Transl Med. 2020 Apr;8(7):501. doi:10.21037/atm.2020.03.63 PubMed PMID: 32395545; PubMed Central PMCID: PMC7210125.

22. Huo B, Boyle A, Marfo N, Tangamornsuksan W, Steen JP, McKechnie T, et al. Large Language Models for Chatbot Health Advice Studies: A Systematic Review. JAMA Netw Open. 2025 Feb 4;8(2):e2457879. doi:10.1001/jamanetworkopen.2024.57879

23. Jafari O, Ma S, Lam BD, Jiang JY, Zhou E, Ranjan M, et al. Development and validation of venous thromboembolism-bidirectional encoder representations from transformers (VTE-BERT) natural language processing model. J Thromb Haemost JTH. 2025 Aug 5;S1538-7836(25)00484–2. doi:10.1016/j.jtha.2025.07.021 PubMed PMID: 40754035; PubMed Central PMCID: PMC12360494.

24. Chiasakul T, Lam BD, McNichol M, Robertson W, Rosovsky RP, Lake L, et al. Artificial intelligence in the prediction of venous thromboembolism: A systematic review and pooled analysis. Eur J Haematol. 2023 Dec;111(6):951–62. doi:10.1111/ejh.14110 PubMed PMID: 37794526; PubMed Central PMCID: PMC10900245.

25. Ma R, Yu W, Tian J, Tang Y, Fang H, Ming X, et al. Machine Learning in the Prediction of Venous Thromboembolism: Systematic Review and Meta-Analysis. J Med Internet Res. 2025 Dec 23;27:e77339. doi:10.2196/77339 PubMed PMID: 41433036; PubMed Central PMCID: PMC12724482.

26. Gu Y, Yang Y, Gao X, Wang Y, Yang L, Wei Y. Application of artificial intelligence in risk assessment and management of venous thromboembolism: scoping review. Front Physiol. 2025;16:1664470. doi:10.3389/fphys.2025.1664470 PubMed PMID: 41132865; PubMed Central PMCID: PMC12540395.

27. Smilg Nicolás C, Tornel Sánchez G, Trujillo Santos J. Concordance among venous thromboembolism risk assessment models. Med Clin (Barc). 2018 Jan 23;150(2):61–3. doi:10.1016/j.medcli.2017.06.021 PubMed PMID: 28743401.

28. Pannucci CJ, Obi A, Alvarez R, Abdullah N, Nackashi A, Hu HM, et al. Inadequate venous thromboembolism risk stratification predicts venous thromboembolic events in surgical intensive care unit patients. J Am Coll Surg. 2014 May;218(5):898–904. doi:10.1016/j.jamcollsurg.2014.01.046 PubMed PMID: 24680577.

29. Merriweather CA, Lyytinen K, Aron D, Cauley MR. When better data meets better design: How EHR data usability and system usability shape physicians’ cognitive load. NPJ Digit Med. 2026 Jan 14;9(1):104. doi:10.1038/s41746-025-02243-4 PubMed PMID: 41535720; PubMed Central PMCID: PMC12864774.

30. Osheroff JA, Teich J, Levick D, Saldana L, Velasco F, Sittig D, et al. Improving Outcomes with Clinical Decision Support: An Implementer’s Guide, Second Edition. 2nd ed. New York: HIMSS Publishing; 2012. 348 p. doi:10.4324/9780367806125

