## Supplementary File for "Multisite Real-World Validation of an Electronic Health Record-Integrated Generative Artificial Intelligence Tool for Venous Thromboembolism Risk Stratification"

### **Appendix A: Johns Hopkins VTE Prevention Program Context**

Johns Hopkins has a long-standing, multidisciplinary, institutional program in venous thromboembolism (VTE) prevention. Early work from the Johns Hopkins multidisciplinary VTE prevention collaborative demonstrated that coordinated, system-level interventions could improve risk-appropriate prophylaxis across hospital settings.^1,2^ Subsequent studies showed that mandatory computerized clinical decision support improved VTE prophylaxis compliance and reduced preventable harm among trauma patients.^3^ Related work evaluating VTE “smart order sets” showed improved prophylaxis compliance and fewer VTE events, supporting order-set design as a modifiable determinant of guideline-concordant care.^4^ Later analyses further demonstrated that mandatory clinical decision support could reduce racial disparities in VTE prophylaxis prescribing, highlighting the relevance of workflow design not only for safety and effectiveness, but also for equitable implementation.^5^

### **Appendix B: Supplementary Methods**

#### **Venous thromboembolism (VTE) Risk Factors and Data Elements**

The IMPROVE guideline for VTE was used as the basis for assessing VTE risk.^1^ Both Johns Hopkins Hospital (JHH) and Bayview Medical Center (BMC) had implemented Johns Hopkins Health System-specific IMPROVE assessment that included: age, history of deep vein thrombosis (DVT) or pulmonary embolism (PE), ICU admission, active cancer, thrombophilia, acute lower limb paralysis, immobility/bedrest, as well as a JHHS-specific list of contraindications to pharmacologic VTE prophylaxis, including high bleeding risk, current anticoagulants, liver disease, activated partial thromboplastin time (aPTT), the international normalized ratio (INR), platelet count, and thrombolytics within 24 hours of ischemic stroke.

#### **Order Sets (Checklist-based vs. Clinician judgement-based)**

Existing clinical workflows at JHHS involve the navigation of order sets (“checklist-based”) for VTE. For each patient, the physician selected risk category (standard, high, or anticoagulated) extracted from VTE prophylaxis order set. Per JHHS protocol, the ‘anticoagulated’ category also belonged to the high-risk category if any of the risk factors was present. A newly developed order set (“clinician judgement-based”) aimed to streamline workflows and clinical practice integration (Appendix A Figure 1). The checklist-based order set workflow designated two risk categories (standard vs. high). The clinician judgement-based order set included “anticoagulated” as a third option in addition to risk selection. Compared to the checklist-based order set, the clinician judgement-based order set retained clinical gestalt as part of the final determination. Among those with clinician judgement-based order set completion, 50 cases were labeled as anticoagulated without specifying high-risk. Nonetheless, receiving anticoagulants reflect elevated risk and is no longer considered as standard risk. Thus, a sensitivity analysis was performed excluding the 50 cases of anticoagulants (binary, standard vs. high-risk) and also mapping those on anticoagulants as high-risk (full, standard vs. high-risk, with anticoagulated as high-risk).

#### **Appendix B Figure S1: New order set (IMPROVE score derived)**

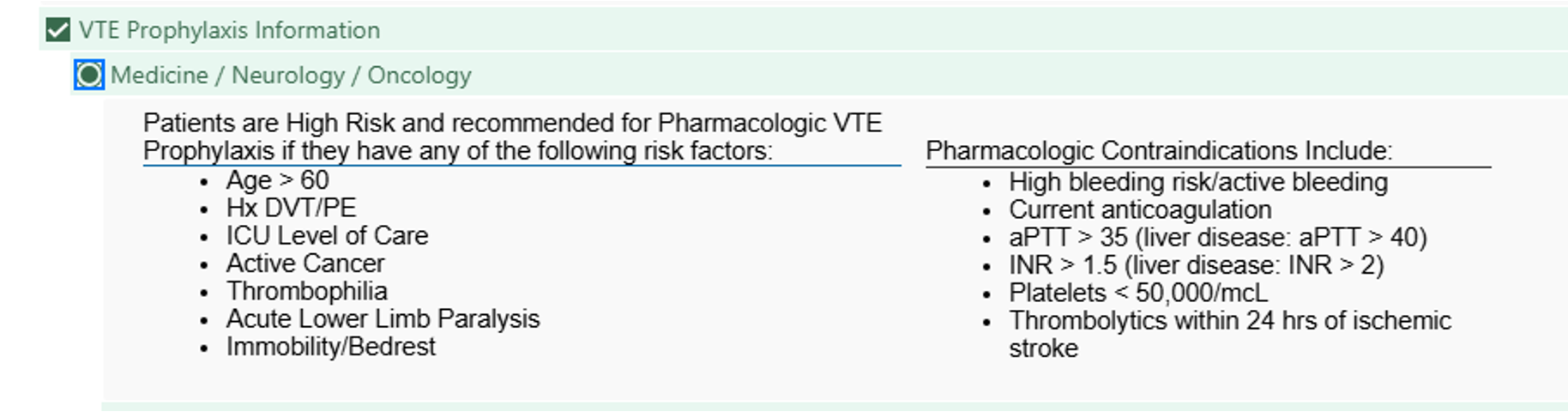

#### **inHealth General Reasoner (iHGR) System**

iHGR is an Epic-integrated GenAI system, developed by the Johns Hopkins inHealth Precision Medicine and Health IT teams in collaboration with Microsoft for automated guideline-based VTE risk classification using EHR data. The system retrieves structured and unstructured patient data through FHIR-based Epic APIs when the patient is admitted, preprocesses these inputs, and applies fixed prompt templates to GPT-4o via an Azure OpenAI service to identify predefined VTE risk factors. Each factor was classified as present or absent with linked supporting source documentation from the EHR. Classified risk factors were then aggregated using predefined guideline-based logic to assign a final binary VTE risk classification (standard vs high-risk).^6^ The workflow, including prompts, extraction logic, inference parameters, and post-processing rules, was maintained under version-controlled operational infrastructure supporting testing, logging, and deployment.

#### **Appendix B Figure S2: Failure modes among iHGR false-negative VTE risk classifications.**

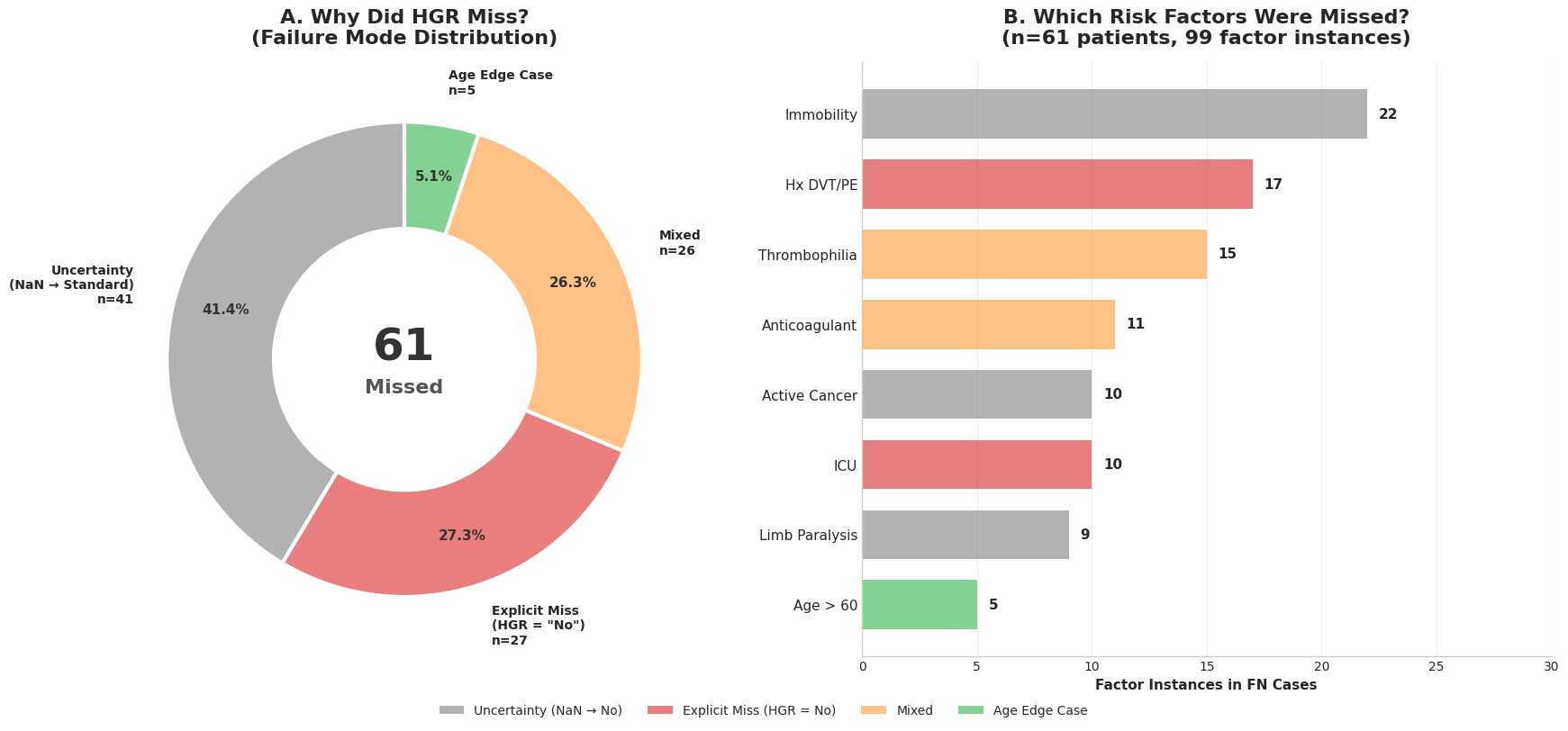

**(a)** Distribution of failure modes among 61 encounters in which iHGR classified the patient as standard risk but physician-adjudicated review classified the patient as high-risk. Failure modes were categorized as uncertainty, explicit miss, mixed, or age edge case. **(b)** Frequency of missed risk-factor instances within false-negative cases. Across 61 patients, 99 missed factor instances were identified; the most common were immobility, history of DVT/PE, thrombophilia, anticoagulant use, active cancer, ICU status, limb paralysis, and age older than 60 years.

### **Appendix C: Supplementary Tables and Figures**

#### **Appendix C Table S1. Clinician Sampling Characteristics by Site and Order Set Period**

| **Metric** | **Value** |
| --- | --- |
| Clinician sample (total unique clinicians) | 75 |
| BMC clinicians | 44 |
| JHH clinicians | 31 |
| Matched clinicians across both periods | 67 (89%) |
| Patients per clinician |  |
| Mean, CHECKLIST-BASED period | 3.5 |
| Mean, CLINICIAN JUDGEMENT-BASED period | 3.6 |
| Median | 3.0 in both periods |
| Range | 1–10 in both periods |

A stratified random sample of 500 patients was drawn from 75 unique clinicians across two sites. To control for clinician-level confounders, only clinicians with patients in both order set periods were prioritized ("matched clinicians"), allowing clinicians to serve as their own controls. A maximum of 8 patients per clinician per stratum was enforced to ensure balanced representation. The similar distribution of patients per clinician across periods (mean 3.5 vs 3.6; median 3.0 vs 3.0) demonstrates successful sampling balance.

#### **Appendix C Figure S1. Clinician Sampling Balance Across Sites and Order Set Periods**

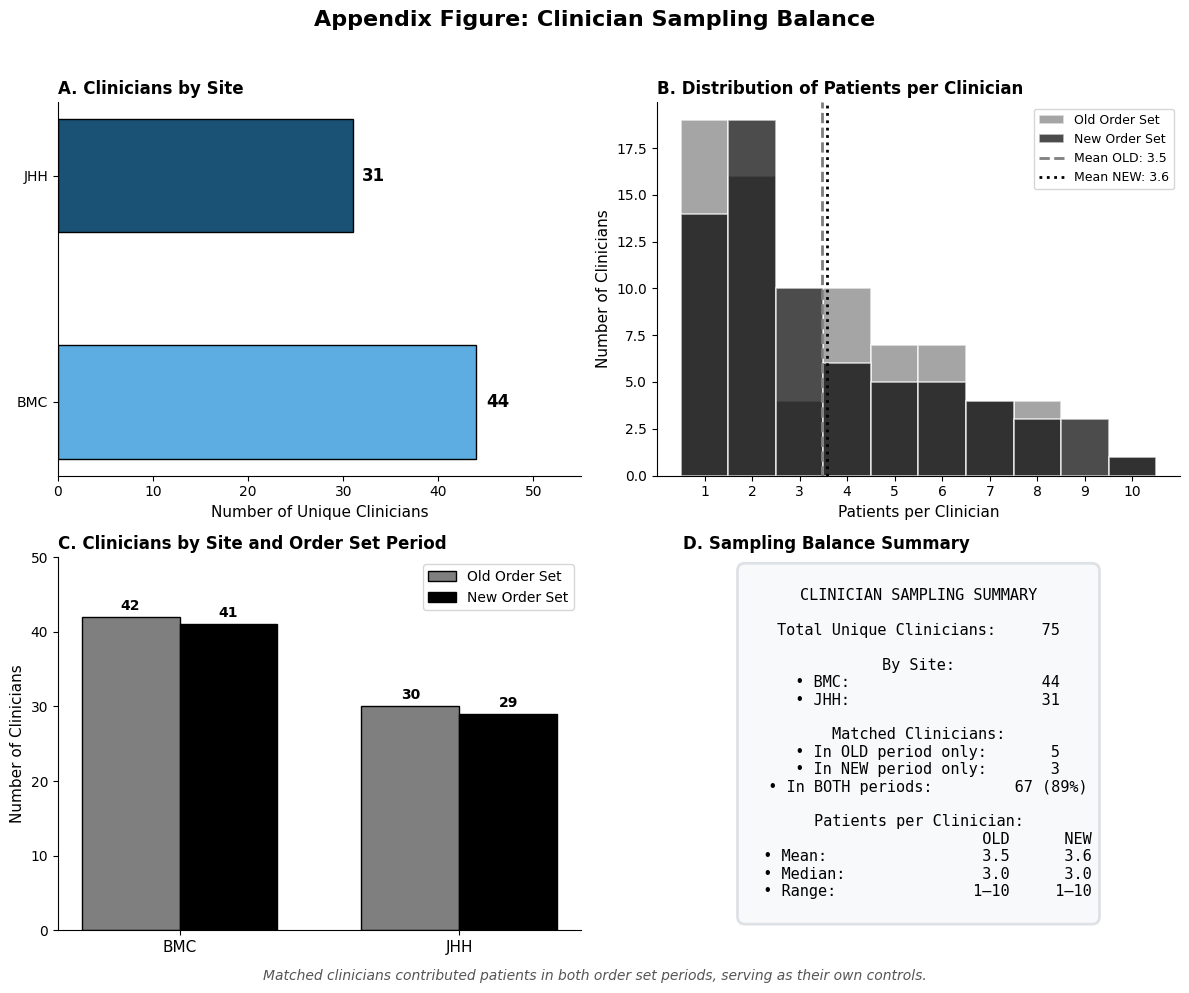

(A) Distribution of unique clinicians by site. A total of 75 clinicians contributed to the analytic sample, with 44 at Bayview Medical Center (BMC) and 31 at Johns Hopkins Hospital (JHH). (B) Histogram of patients per clinician stratified by order set period. The distributions are nearly identical (checklist-based order set: mean 3.5, median 3.0; clinician judgement-based order set: mean 3.6, median 3.0), with both ranging from 1 to 10 patients per clinician. Dashed and dotted vertical lines indicate period-specific means. (C) Clinician counts by site and order set period. Each site contributed similar numbers of clinicians to both the old and clinician judgement-based order set periods, reflecting balanced sampling across strata. (D) Summary of clinician sampling metrics. Of 75 total clinicians, 67 (89%) contributed patients in both order set periods, serving as their own controls for within-clinician comparisons across the workflow change.

#### **Appendix C Table S2. Patient demographics for the analytic sample (n=500) stratified by site.**

|  | **Analytic Sample** | **BMC** | **JHH** | **P-Value** |
| --- | --- | --- | --- | --- |
| **N** | 500 | 250 | 250 |  |
| Age, median [Q1,Q3] | 63.0 [46.8,75.6] | 67.5 [49.9,78.4] | 60.2 [44.2,72.2] | <0.001 |
| Sex, n (%) |  |  |  |  |
| Female | 275 (55.0) | 146 (58.4) | 129 (51.6) | 0.150 |
| Male | 225 (45.0) | 104 (41.6) | 121 (48.4) |  |
| Race, n (%) |  |  |  |  |
| White | 257 (51.4) | 161 (64.4) | 96 (38.4) | <0.001 |
| Black | 201 (40.2) | 72 (28.8) | 129 (51.6) |  |
| Other/Multiple | 29 (5.8) | 14 (5.6) | 15 (6.0) |  |
| Asian | 12 (2.4) | 3 (1.2) | 9 (3.6) |  |
| Unknown/Not Disclosed | 1 (0.2) |  | 1 (0.4) |  |
| Ethnicity, n (%) |  |  |  |  |
| Not Hispanic/Latino | 369 (73.8) | 196 (78.4) | 173 (69.2) | 0.062 |
| Other/Unknown | 113 (22.6) | 46 (18.4) | 67 (26.8) |  |
| Hispanic/Latino | 18 (3.6) | 8 (3.2) | 10 (4.0) |  |
| Marital Status, n (%) |  |  |  |  |
| Married | 149 (29.8) | 68 (27.2) | 81 (32.4) | <0.001 |
| Single | 208 (41.6) | 89 (35.6) | 119 (47.6) |  |
| Widowed | 69 (13.8) | 48 (19.2) | 21 (8.4) |  |
| Divorced | 53 (10.6) | 29 (11.6) | 24 (9.6) |  |
| Other/Unknown | 21 (4.2) | 16 (6.4) | 5 (2.0) |  |
| Employment, n (%) |  |  |  |  |
| Retired | 195 (39.0) | 114 (45.6) | 81 (32.4) | 0.003 |
| Employed | 103 (20.6) | 39 (15.6) | 64 (25.6) |  |
| Not Employed | 131 (26.2) | 68 (27.2) | 63 (25.2) |  |
| Disabled | 59 (11.8) | 22 (8.8) | 37 (14.8) |  |
| Other/Unknown | 12 (2.4) | 7 (2.8) | 5 (2.0) |  |
| Language, n (%) |  |  |  |  |
| English | 474 (94.8) | 241 (96.4) | 233 (93.2) | 0.272 |
| Spanish | 6 (1.2) | 2 (0.8) | 4 (1.6) |  |
| Other | 20 (4.0) | 7 (2.8) | 13 (5.2) |  |
| Charlson Comorbidity Score, median [Q1,Q3] | 2.0 [0.0,3.0] | 2.0 [1.0,3.0] | 2.0 [0.0,3.0] | 0.850 |
| Length of Stay (days), median [Q1,Q3] | 4.0 [2.0,8.0] | 4.0 [2.0,8.0] | 5.0 [3.0,9.0] | 0.085 |
| Mortality, n (%) | 30 (6.0) | 14 (5.6) | 16 (6.4) | 0.851 |

#### **Appendix C Table S3. Inter-rater reliability between reviewers.**

| **Metric** | **Reviewer 1 vs. Reviewer 2** |
| --- | --- |
| Basic Agreement |  |
| N (Both Reviewed) | 500 |
| N Agreement | 378 |
| % Agreement | 75.6% |
| Chance-Corrected |  |
| Cohen's Kappa | 0.488 [0.409, 0.567] |
| PABAK | 0.512 [0.433, 0.583] |
| Gwet's AC1 | 0.534 [0.458, 0.609] |
| Krippendorff's Alpha | 0.489 [0.410, 0.567] |
| Directional Agreement |  |
| Prevalence of High-risk | 0.608 |
| High-risk Agreement | 0.799 [0.766, 0.829] |
| Standard Risk Agreement | 0.689 [0.641, 0.733] |
| Symmetry Test |  |
| McNemar's χ² | 0 |
| McNemar's p-value | 1 |

#### **Appendix C Table S4. GS chart review variables stratified by iHGR classification categories.**

| **GS Chart Review Variables** | **Overall** | **iHGR vs. GS Chart Review** | | | |
| --- | --- | --- | --- | --- | --- |
|  | **(N=500)** | **FN (N=61)** | **FP (N=48)** | **TN (N=117)** | **TP (N=274)** |
| High-risk - Active Cancer | 6 (1.2) | 0 (0.0) | 0 (0.0) | 0 (0.0) | 6 (2.2) |
| High-risk - Active Cancer, Acute Limb Paralysis, Age > 60, History of VTE (e.g., DVT, PE) | 2 (0.4) | 0 (0.0) | 0 (0.0) | 0 (0.0) | 2 (0.7) |
| High-risk - Active Cancer, Age > 60 | 15 (3.0) | 0 (0.0) | 1 (2.1) | 0 (0.0) | 14 (5.1) |
| High-risk - Active Cancer, Age > 60, History of VTE (e.g., DVT, PE) | 11 (2.2) | 0 (0.0) | 0 (0.0) | 0 (0.0) | 11 (4.0) |
| High-risk - Active Cancer, Age > 60, History of VTE (e.g., DVT, PE), Immobility/Bed Rest, Thrombophilia | 1 (0.2) | 0 (0.0) | 0 (0.0) | 0 (0.0) | 1 (0.4) |
| High-risk - Active Cancer, Age > 60, ICU Level of Care | 1 (0.2) | 0 (0.0) | 0 (0.0) | 0 (0.0) | 1 (0.4) |
| High-risk - Active Cancer, Age > 60, Immobility/Bed Rest | 1 (0.2) | 0 (0.0) | 0 (0.0) | 0 (0.0) | 1 (0.4) |
| High-risk - Active Cancer, History of VTE (e.g., DVT, PE) | 4 (0.8) | 0 (0.0) | 0 (0.0) | 0 (0.0) | 4 (1.5) |
| High-risk - Active Cancer, History of VTE (e.g., DVT, PE), Immobility/Bed Rest | 3 (0.6) | 0 (0.0) | 0 (0.0) | 0 (0.0) | 3 (1.1) |
| High-risk - Active Cancer, Immobility/Bed Rest | 1 (0.2) | 0 (0.0) | 0 (0.0) | 0 (0.0) | 1 (0.4) |
| High-risk - Acute Limb Paralysis | 1 (0.2) | 0 (0.0) | 1 (2.1) | 0 (0.0) | 0 (0.0) |
| High-risk - Acute Limb Paralysis, Age > 60, History of VTE (e.g., DVT, PE) | 1 (0.2) | 0 (0.0) | 0 (0.0) | 0 (0.0) | 1 (0.4) |
| High-risk - Acute Limb Paralysis, Age > 60, Immobility/Bed Rest | 1 (0.2) | 0 (0.0) | 0 (0.0) | 0 (0.0) | 1 (0.4) |
| High-risk - Age > 60 | 177 (35.4) | 0 (0.0) | 38 (79.2) | 0 (0.0) | 139 (50.7) |
| High-risk - Age > 60, History of VTE (e.g., DVT, PE) | 56 (11.2) | 0 (0.0) | 1 (2.1) | 0 (0.0) | 55 (20.1) |
| High-risk - Age > 60, History of VTE (e.g., DVT, PE), Thrombophilia | 1 (0.2) | 0 (0.0) | 0 (0.0) | 0 (0.0) | 1 (0.4) |
| High-risk - Age > 60, Immobility/Bed Rest | 2 (0.4) | 0 (0.0) | 2 (4.2) | 0 (0.0) | 0 (0.0) |
| High-risk - History of VTE (e.g., DVT, PE) | 34 (6.8) | 0 (0.0) | 5 (10.4) | 0 (0.0) | 29 (10.6) |
| High-risk - History of VTE (e.g., DVT, PE), ICU Level of Care, Immobility/Bed Rest | 1 (0.2) | 0 (0.0) | 0 (0.0) | 0 (0.0) | 1 (0.4) |
| High-risk - History of VTE (e.g., DVT, PE), Immobility/Bed Rest | 1 (0.2) | 0 (0.0) | 0 (0.0) | 0 (0.0) | 1 (0.4) |
| High-risk - History of VTE (e.g., DVT, PE), Thrombophilia | 1 (0.2) | 0 (0.0) | 0 (0.0) | 0 (0.0) | 1 (0.4) |
| High-risk - Thrombophilia | 1 (0.2) | 0 (0.0) | 0 (0.0) | 0 (0.0) | 1 (0.4) |
| Standard Risk | 178 (35.6) | 61 (100.0) | 0 (0.0) | 117 (100.0) | 0 (0.0) |
| iHGR: inHealth General Reasoner. GS: Gold standard chart review. TP: True positive. FP: False positive. FN: False negative. TN: True negative. | | | | | |

### **APPENDIX REFERENCES**

1. Lau BD, Haut ER. Practices to prevent venous thromboembolism: a brief review. BMJ Qual Saf. 2014 Mar;23(3):187–95. doi:[10.1136/bmjqs-2012-001782](https://doi.org/10.1136/bmjqs-2012-001782) PubMed PMID: 23708438; PubMed Central PMCID: PMC3932749.
2. Streiff MB, Carolan HT, Hobson DB, Kraus PS, Holzmueller CG, Demski R, et al. Lessons from the Johns Hopkins Multi-Disciplinary Venous Thromboembolism (VTE) Prevention Collaborative. BMJ. 2012 Jun 19;344:e3935. doi:[10.1136/bmj.e3935](https://doi.org/10.1136/bmj.e3935) PubMed PMID: 22718994; PubMed Central PMCID: PMC4688421.
3. Haut ER, Lau BD, Kraenzlin FS, Hobson DB, Kraus PS, Carolan HT, et al. Improved prophylaxis and decreased rates of preventable harm with the use of a mandatory computerized clinical decision support tool for prophylaxis for venous thromboembolism in trauma. Arch Surg. 2012 Oct;147(10):901–7. doi:[10.1001/archsurg.2012.2024](https://doi.org/10.1001/archsurg.2012.2024) PubMed PMID: 23070407.
4. Zeidan AM, Streiff MB, Lau BD, Ahmed SR, Kraus PS, Hobson DB, et al. Impact of a venous thromboembolism prophylaxis “smart order set”: Improved compliance, fewer events. Am J Hematol. 2013 Jul;88(7):545–9. doi:[10.1002/ajh.23450](https://doi.org/10.1002/ajh.23450) PubMed PMID: 23553743.
5. Lau BD, Haider AH, Streiff MB, Lehmann CU, Kraus PS, Hobson DB, et al. Eliminating Health Care Disparities With Mandatory Clinical Decision Support: The Venous Thromboembolism (VTE) Example. Med Care. 2015 Jan;53(1):18–24. doi:[10.1097/MLR.0000000000000251](https://doi.org/10.1097/MLR.0000000000000251) PubMed PMID: 25373403; PubMed Central PMCID: PMC4262632.
6. Rosenberg D, Eichorn A, Alarcon M, McCullagh L, McGinn T, Spyropoulos AC. External validation of the risk assessment model of the International Medical Prevention Registry on Venous Thromboembolism (IMPROVE) for medical patients in a tertiary health system. J Am Heart Assoc. 2014 Nov 17;3(6):e001152. doi:[10.1161/JAHA.114.001152](https://doi.org/10.1161/JAHA.114.001152) PubMed PMID: 25404191; PubMed Central PMCID: PMC4338701.
